# Systematic Review of Genetic Polymorphisms Associated with Acute Pain and Relevant Acute Toxicities Induced by Radiotherapy for Head and Neck Cancer

**DOI:** 10.1101/2022.11.15.22282359

**Authors:** Vivian Salama, Yimin Geng, Jillian Rigert, Clifton D. Fuller, Amy C. Moreno, Sanjay Shete, Cielito C. Reyes-Gibby

**Author notes:** Corresponding authors Vivian Salama;, Cielito C. Reyes-Gibby.

## Abstract

**Background/objective:** Pain is the most common acute toxicity following radiation therapy (RT) for head and neck cancer (HNC). The multifactorial origin of radiotherapy-induced pain makes it highly challenging to manage in HNC patients. Multiple studies have been conducted to identify different germline genetic variants associated with cancer pain, however few of them focused on RT-induced acute pain. In this systematic review, we summarize potential mechanisms of acute pain after radiotherapy in HNC focusing on oral cavity/oropharyngeal cancer and identify genetic variants associated with radiotherapy-induced acute pain and other relevant acute toxicities.

**Methods:** A comprehensive search of Ovid Medline, EMBASE and Web of Science databases using concepts and terms including “Variants”, “Polymorphisms”, “Radiotherapy”, “Acute pain”, “Acute toxicity” published up to February 28, 2022 was performed by two reviewers. Review articles and citations were reviewed manually. The reported SNPs associated with RT-induced acute pain and toxicities were reported, and the molecular function of the associated genes and pathways were described based on genetic annotation using The Human Gene Database; GeneCards.

**Results:** A total of 386 articles were identified electronically and an additional 8 articles were included after manual search. 39 articles were finally included. 51 variants were associated with 40 genes, of which 30 % had function in DNA damage response and repair, 25% in inflammatory and immune response, 17.5 % in cell death or cell cycle, and were associated with RT-inflammatory pain and acute mucositis or dermatitis. 4 variants in 4 genes were associated with neuropathy and neuropathic pain. 13 variants in 10 genes and were associated with RT-induced mixed types of post-RT-pain.

**Conclusion:** Different types of pain develop after RT, including inflammatory pain (acute mucositis and acute skin reaction); neuropathic pain; nociceptive pain; and mixed oral pain. Genetic variants involved in DNA damage response and repair, cell death, inflammation and neuropathic pathways may affect pain presentation post-RT. These variants could be used for acute pain prediction and personalized pain management in HNC patients receiving RT.

## Introduction

Head and neck squamous cell carcinoma (HNSCC) are composed of a heterogenous mix of malignant epithelial cancers that arises from the oral cavity, pharynx or larynx. Oral cavity and oropharyngeal (OC/OPC) squamous cell carcinoma, afflicted more than 54,000 patients in the U.S, in 2022 ^1^.Management of OC/OPC is a stage-dependent multimodality approach that entails surgery, chemotherapy, and radiotherapy. Radiotherapy (RT) is a cornerstone for the management of OC/OPC patients. RT, especially after application of the novel RT techniques, has a favorable impact on the overall survival rates ^2^. Despite improvement in survival rates, various significant acute and sometimes chronic or permanent toxicities are reported during and after RT. RT-attributable toxicities, including mucositis, dermatitis and dysphagia, have significant negative impacts on patients’ quality of life (QOL) and may rarely even lead to death ^3–5^.

Oral/throat pain, the most common radiation-attributable acute toxicity during and after locoregional RT of OC/OPC, is associated with increase in analgesics use ^6^. About 68-86% of OC/OPC patients and about one-third of HNC patients present to the emergency department with uncontrolled pain ^7–11^. The control of RT-induced acute pain is challenging because of the complex nature and multiple mechanisms of pain in these patients. Baseline pain and associated RT toxicities such as mucositis, dermatitis, neuropathy, and dysphagia are considered the most common etiologies of RT-induced pain. In addition, pain due to surgery and/or chemotherapy are also observed in these patients ^12–14^. Most clinicians will prescribe opioids to OC/OPC patients during cancer therapy, with 15-40% of patients dependent on opioids for several months post therapy ^15–17^. The optimal use of opioids is a major clinical challenge. While trying not to undertreat patients with severe pain, potential overuse of opioids may negatively impact the health status and the quality of life of patients. Furthermore, for a subset of patients, the prolonged use of opioids increases the risk for drug abuse and addiction, which may negatively impact the QOL and increase the potential for overdose and death ^18^.

### The mechanism of radiotherapy-induced cell death and normal tissue toxicities

<u>Radiation therapy</u> is a group of ionizing energy beams that induce DNA damage in cells, cancer cells being most susceptible due to their characteristic of rapid cell division. Direct DNA damage is induced by ionizing radiation to DNA, inducing DNA breaks (single-strand breaks (SSBs), double strand breaks (DSBs)) and covalent crosslinking of the complementary DNA strands ^19, 20^. Indirect DNA damage occurs through RT-induced generation of free radicals. For instance, reactive oxygen species (ROS) are produced by the interaction of ionizing radiation with the water molecules ^21^. After RT, the impairment of DNA repair mechanisms in cancer cells in addition to the accumulation of intracellular ROS induces cell injury and apoptosis, necrosis and cellular senescence ^22–25^. However, normal neighboring healthy tissues will also be exposed to ionizing radiation, inducing normal cell injury and maybe cell death despite the lower doses of radiation exposure. Normal tissue damage induces toxicities such as oral mucositis, dermatitis, and neuropathy leading to aggravation of RT-induced acute pain. The severity of RT-induced toxicities and pain is dependent on the total dose, fractions and the duration of radiation delivered to normal tissue in addition to the variation in tissue tolerance to radiation ^26, 27^.

### Types and potential mechanisms of radiotherapy induced acute pain in OC/OPC patients

Adequate management of acute pain in OC/OPC patients during RT requires good understanding of the different types and mechanisms of RT-induced pain. Three main types of RT-induced acute pain have been identified in HNC patients receiving RT ^28–30^:

1. Inflammatory pain
2. Nociceptive pain
3. Neuropathic pain

#### 1. Inflammatory pain

Inflammatory pain, the most common type of RT-induced acute pain in HNC, is caused by activation of inflammatory and immune cells in response to tissue injury and infections induced by RT ^21, 31^. Inflammatory and immune cells in addition to RT-induced damaged cells release inflammatory mediators such as pro-inflammatory cytokines: interleukin (IL)-1β, IL-6, NF-kB, prostaglandins (PG), and tumor necrosis factor-α (TNF-α) ^21, 32, 33^. These cytokines induce more recruitment of immune and inflammatory cells, inducing more damage of tissues ^34^. Oral/pharyngeal mucositis and neck dermatitis are the most common inflammatory reactions induced by RT ^21, 33, 35, 36^. Different signaling pathways involved in RT-induced mucositis and dermatitis have been identified, and they may contribute to RT-induced inflammatory pain, such as: nuclear factor-κB (NF-κB) signaling which upregulates other pathways such as COX2 pathway and downstream tyrosine kinase receptor pathways (e.g. PI3K/AKT signaling and MAPK signaling). Other signaling pathways such as DNA damage checkpoints, cell cycle pathways, WNT/B-catenin and integrin signaling, VEGF signaling, glutamate receptor signaling and IL-6 signaling, have also been identified in correlation with RT-induced mucositis ^36, 37^.

#### 2. Nociceptive pain

Nociceptive pain is caused by direct damage to non-neural tissue, often caused by an external injury which stimulates nociceptive receptors ^30^. Noxious stimuli are released from either damaged cancer, normal cells or from inflammatory/immune cells recruited after RT ^38^. Noxious stimuli activate the peripheral sensory neurons through stimulation of afferent sensory neurons which transmit the action potential to neuronal bodies where calcium influx occurs, leading to the release of neurotransmitters (e.g.: substance-P, glutamate, γ-aminobutyric acid (GABA), adenosine triphosphate (ATP), glycine, dopamine, norepinephrine, nitric oxide, and serotonin) which bind to the post-synaptic membrane receptors. The signal then reaches the second order neuron and is transmitted to the somatosensory cortex of the brain where the pain is perceived ^39–43^. The trigeminal nerves and the facial nerves (CN VII) play important roles in pain perception in the head and neck ^44^. RT-damaged cells and infiltrating immune cells release multiple noxious stimuli. Cytokines and inflammatory mediators released from infiltrating immune cells such as IL-1, TNF-α and IL-6 induce nociceptor sensitization. In addition, the acidic pH of the tumor microenvironment and extracellular ATP act as noxious stimuli to nociceptors at the cancer site ^45^.

#### 3. Neuropathic pain

Neuropathic pain is caused by damage to nerves or nervous system ^46^. RT directly injures the somatosensory nervous system promoting pain signal transmission and peripheral neuropathic pain. RT induces DNA damage which induces apoptosis. Apoptosis is induced through p53 activation which activates the cascade to execute cell death ^47, 48^. Endothelial cell death reduces blood flow to peripheral nerves, including damage and eventual neuronal fibrosis. Furthermore, ionizing radiation and ROS released after RT, cause neuronal cell stress and direct nerve damage. In RT of HNC, the brachial plexus are the neural tissues at high risk of damage. Overexpression of p53 induces apoptosis of neurons after radiation exposure ^49^. Furthermore, RT induces inflammation and infiltration by immune cells, resulting in immune-mediated peripheral neuropathy. Chronic inflammation in the nervous system microenvironment promotes neuron loss with fibrosis, resulting in chronic neuropathic pain ^50^.

### Genetic variants associated with radiotherapy induced acute pain

Advances in molecular and genetic technologies have motivated researchers and scientists toward exploring the human genome and analyzing genetic variations and their correlations with different diseases and treatment outcome.

RT-induced acute pain presents a significant morbidity burden on OC/OPC patients receiving RT and drastically reduces patients’ quality of life. In an effort to better predict and optimize pain management approaches, various cellular and molecular approaches have been widely explored recently to identify the genomic biomarkers for patients vulnerable to develop RT-induced pain, and the influence of these genetic variants on pain modulation and analgesic response ^51, 52^. The advanced technologies in human genome sequencing allowed deep understanding of the genetic variations and mutations related to cancer treatment associated pain. Single nucleotide polymorphisms (SNPs) are the most common DNA sequence variations. These genetic polymorphisms are stable markers and easily and reliably assayed to explore the extent to which genetic variation might prove useful in identifying patients with cancer at high-risk of pain development and their response to pain therapies ^53^. Likewise, these candidate SNPs could be used in building robust predictive models for pre-treatment prediction of acute as well as chronic toxicities for personalized management and precision medicine ^54^.

Given the multifactorial and complex etiology of RT-induced pain in HNC patients, the challenges in managing RT-associated pain, and the adverse impact of pain on patients’ quality of life, the aim of this literature review is to identify predictive genetic variants and pathways associated with RT-induced pain and related phenotypes. This synthesis of the current literature will use provide the basis to develop predictive models of RT-induced acute pain to assist with personalized analgesic therapy.

## Materials and Methods

### Search Strategies

We conducted a systematic search of databases including Ovid MEDLINE, Ovid Embase, and Clarivate Analytics Web of Science, for publications in English language from the inception of databases to February 28, 2022. The following concepts were searched using subject headings and free text keywords as needed, “radiotherapy”, “radiation therapy”, “pain”, “neuropathy”, “analgesics”, “opioid”, “acute toxicity”, “mucositis”, “dermatitis”, “single nucleotide polymorphism”, “genetic variation”, “genetic variability”, and “genetic predisposition” etc. The terms were combined using AND/OR Boolean Operators. Animal studies, in vitro studies, and conference abstracts were removed from the search result. Handsearching of journals and publisher databases and manually checking the reference lists of journal articles were also conducted to supplement the electronic database search. The complete electronic database search strategies were detailed in Supplementary Tables S1–S3.

### Inclusion criteria

The included studies met all of the following inclusion criteria: 1) articles published in English, 2) human studies in any type of cancers, 3) genetic polymorphisms reported to be significantly associated with RT-induced pain in different phenotypes (Inflammatory [Mucositis, dermatitis], neuropathy, nociceptive pain or mixed oral/throat pain). We included variants associated with our phenotypes in any type of cancer, for comprehensive search intent and to identify new SNPs identified in other cancers that need more focus in HNC.

### Exclusion criteria

Articles were excluded if they met any of the following criteria: 1) non-genetic article or no genetic association study, 2) not radiotherapy induced pain or toxicity, 3) meta-analysis, or review article, 4) articles written in languages other than English, 5) clinical trials, 6) non-human study, or not blood or buccal DNA or 7) Other unrelated phenotypes like dysphagia, gastrointestinal toxicities (e.g. diarrhea).

### Functional annotation

Manual functional annotation of the identified genes was done using GeneCards database. The function of each gene was assigned according to the most common functional pathway annotated in the GenCards and the relevant studies published online. ^55, 56^ We divided the collected variants and genes into three groups: 1) variants associated with RT-induced inflammatory pain including; RT-induced acute toxicities; acute mucositis and acute dermatitis (or acute skin reaction), 2) variants associated with other types of RT-induced acute pain (including post RT-pain, post-RT throat/mouth/neck pain and neuropathic pain/neuropathy)

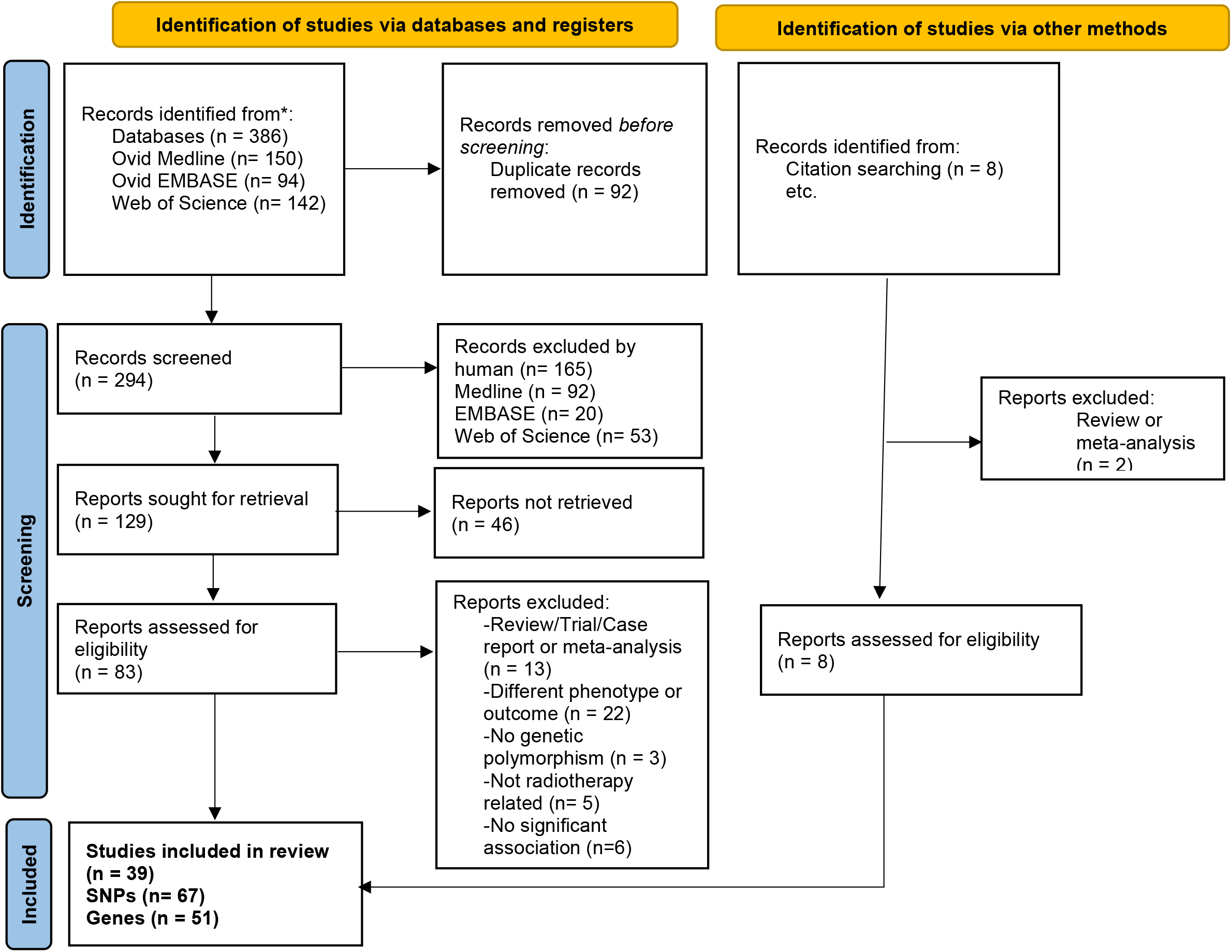
Flow Chart of systematic review of SNPs associated Radiation-induced acute pain and relevant acute toxicities.

## Results

### 1 Variants associated with radiotherapy induced inflammatory pain

Mucositis and acute skin reactions are the most common acute reactions reported following RT. 51 variants in 40 genes were associated with radiotherapy induced inflammatory pain and acute reactions including mucositis or dermatitis. (Table 1). Table 2 shows the biological functions and pathways of the genes associated with RT-induced acute mucositis and acute skin reaction. We found that 12 genes (32%) were involved in DNA damage and DNA repair pathways. 10 genes (19.5%) were involved in inflammatory pathways and immune systems, 7 genes (19.5%) were regulating cell death, apoptosis, autophagy or cell cycle pathways, 6 genes (17%) were involved in metabolisms and microenvironment functional pathways, 5 genes were functionally involved in other functional pathways. (Figure 2, table 2)

**Table 1:**
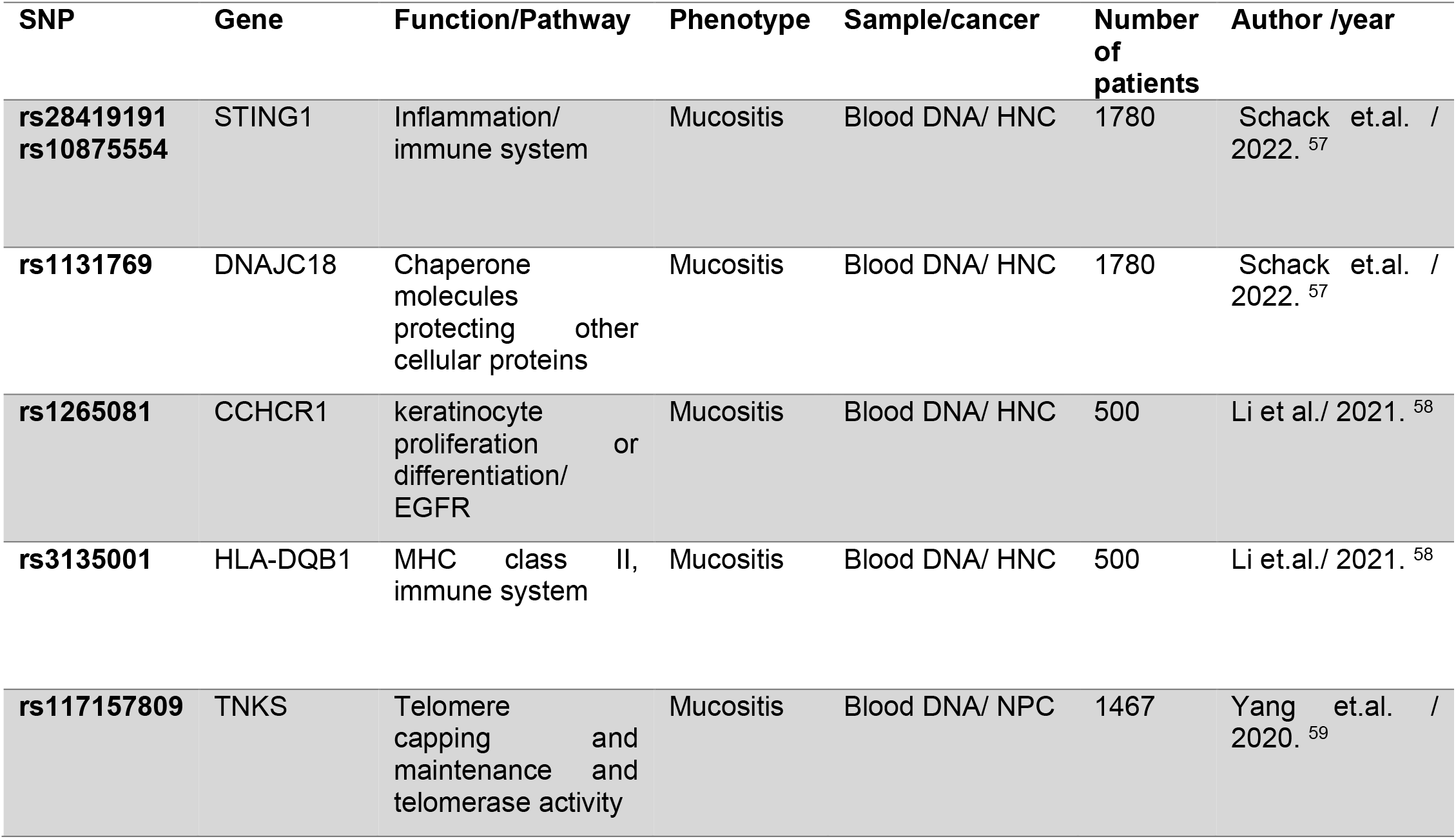
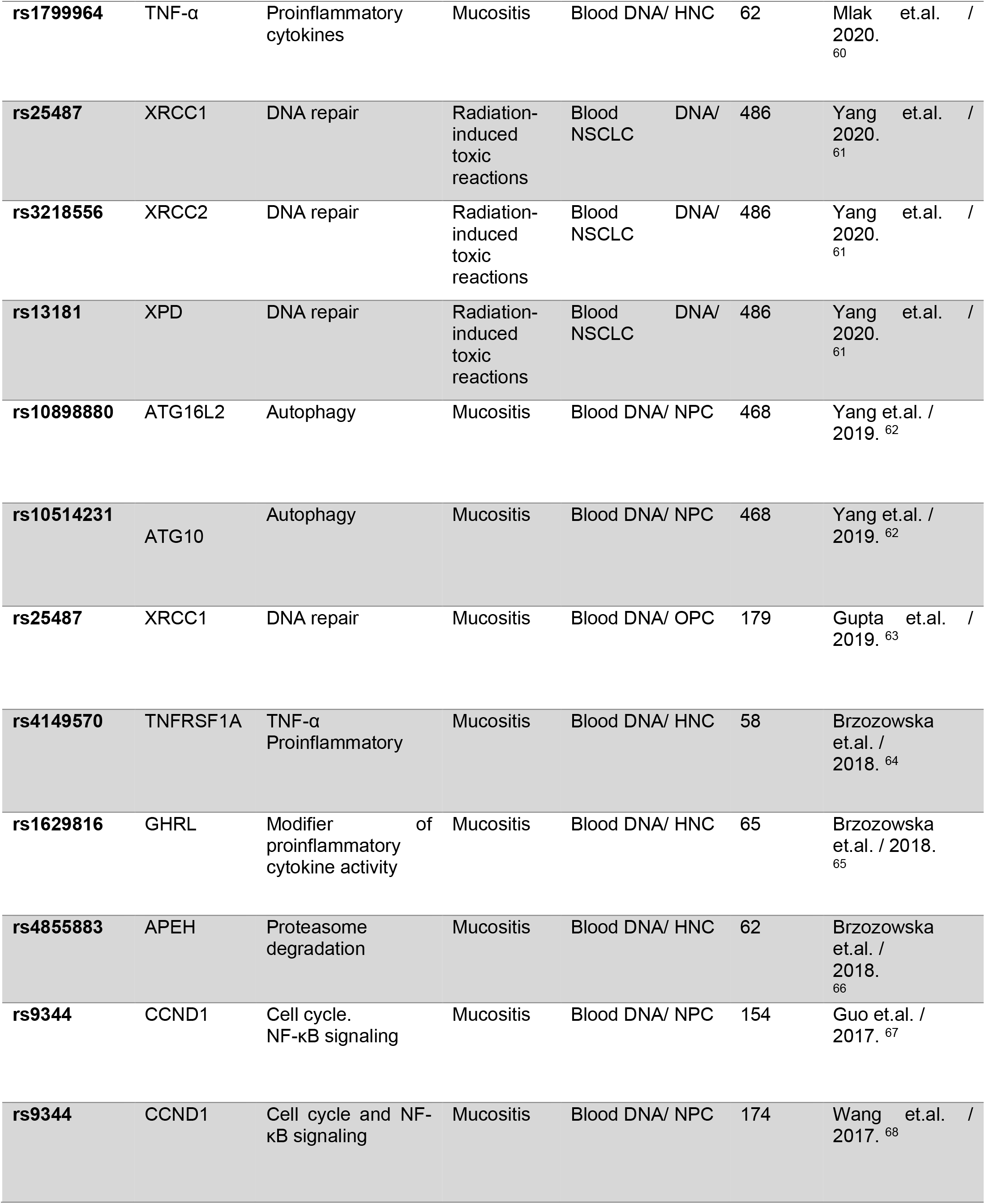
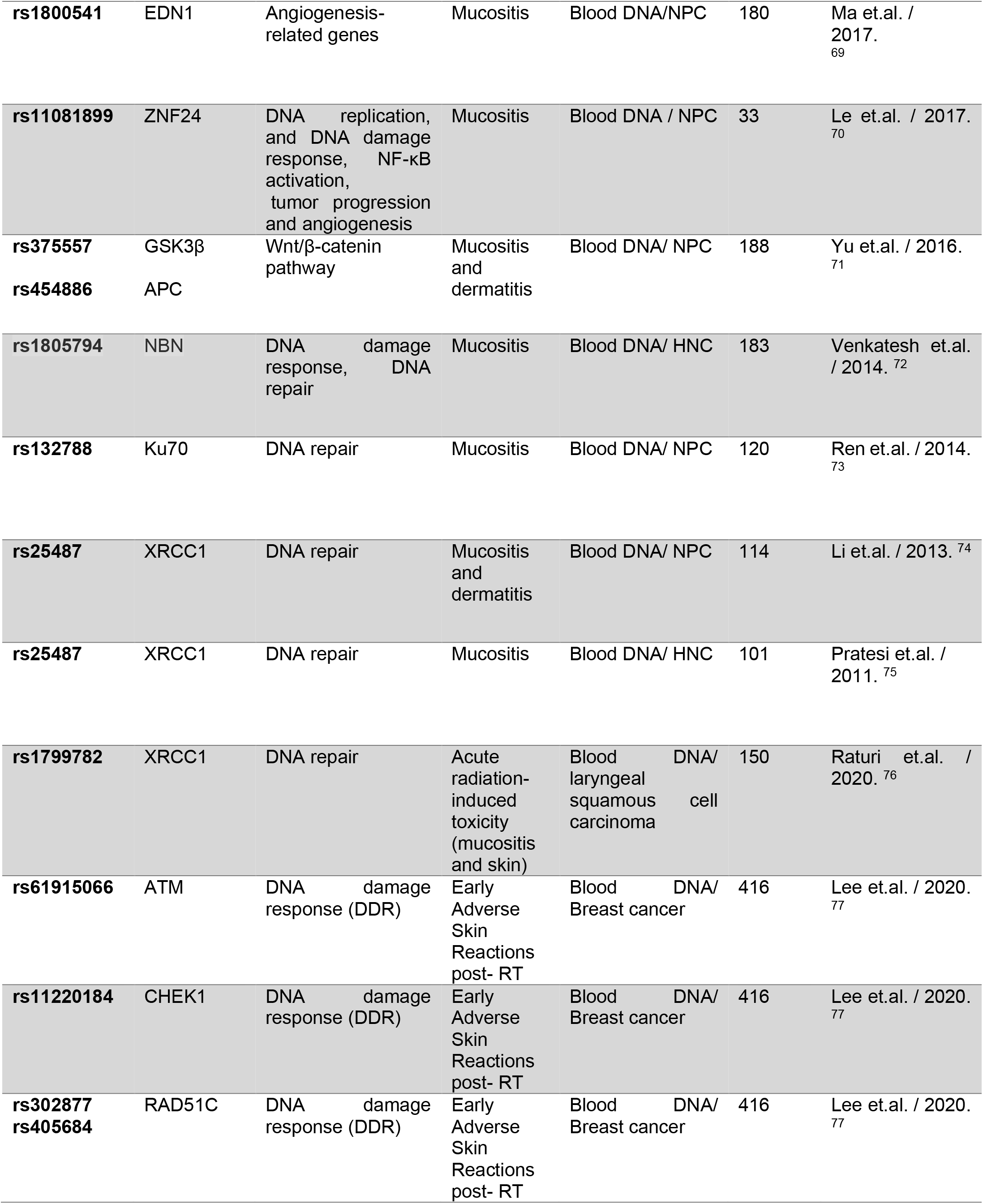
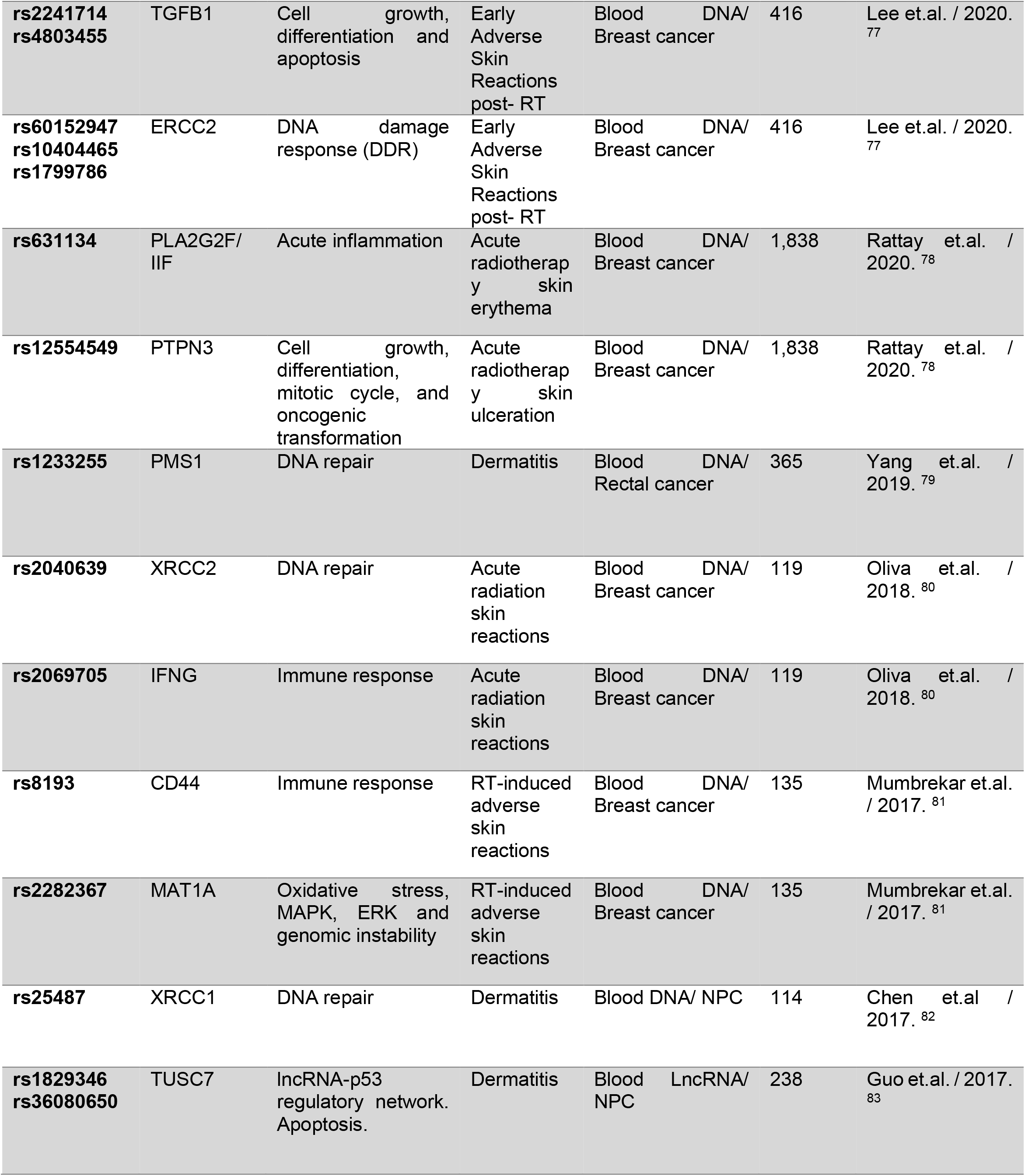
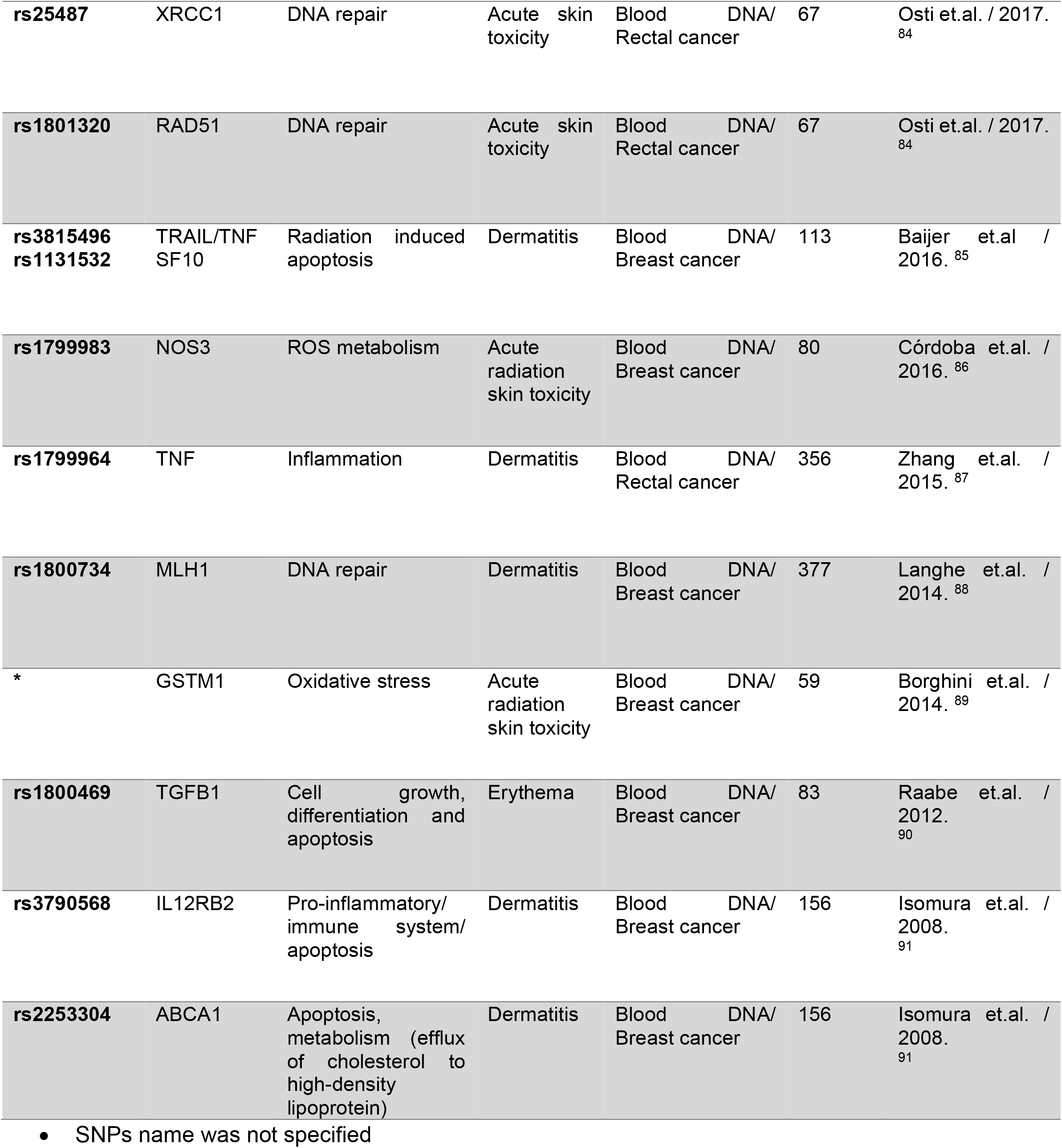
Summary of literature search of variants associated with radiotherapy induced acute inflammatory pain.

**Table 2:** Pathways and biological functions of genes associated with radiotherapy induced acute inflammatory pain.

| Pathway | Genes | Number (n) | Percent (%) |
| --- | --- | --- | --- |
| **DNA damage / DNA repair** | XRCC1, XRCC2, XPD, ZNF24, NBN, Ku70, ATM, CHEK1, RAD51C, ERCC2, PMS1, MLH1 | 12 | 30 % |
| **Inflammatory/ Immune system** | STING1, HLA-DQB1, TNF-α, TNFRSF1A, GHRL, PLA2G2F/ IIF, IFNG, IL12RB2, CD44, TNF | 10 | 25 % |
| **Cell death/ Apoptosis/Autophagy/ Cell cycle** | ATG16L2, ATG10, TUSC7, ABCA1, CCND1, TGFB1, TRAIL/TNFSF10 | 7 | 17.5 % |
| **Metabolism/ Microenvironment** | APEH, EDN1, GSK3β, APC, NOS3, GSTM1 | 6 | 15 % |
| **Others** | DNAJC18, CCHCR1, TNKS, PTPN3, MAT1A | 5 | 12.5 % |

**Figure 1:**
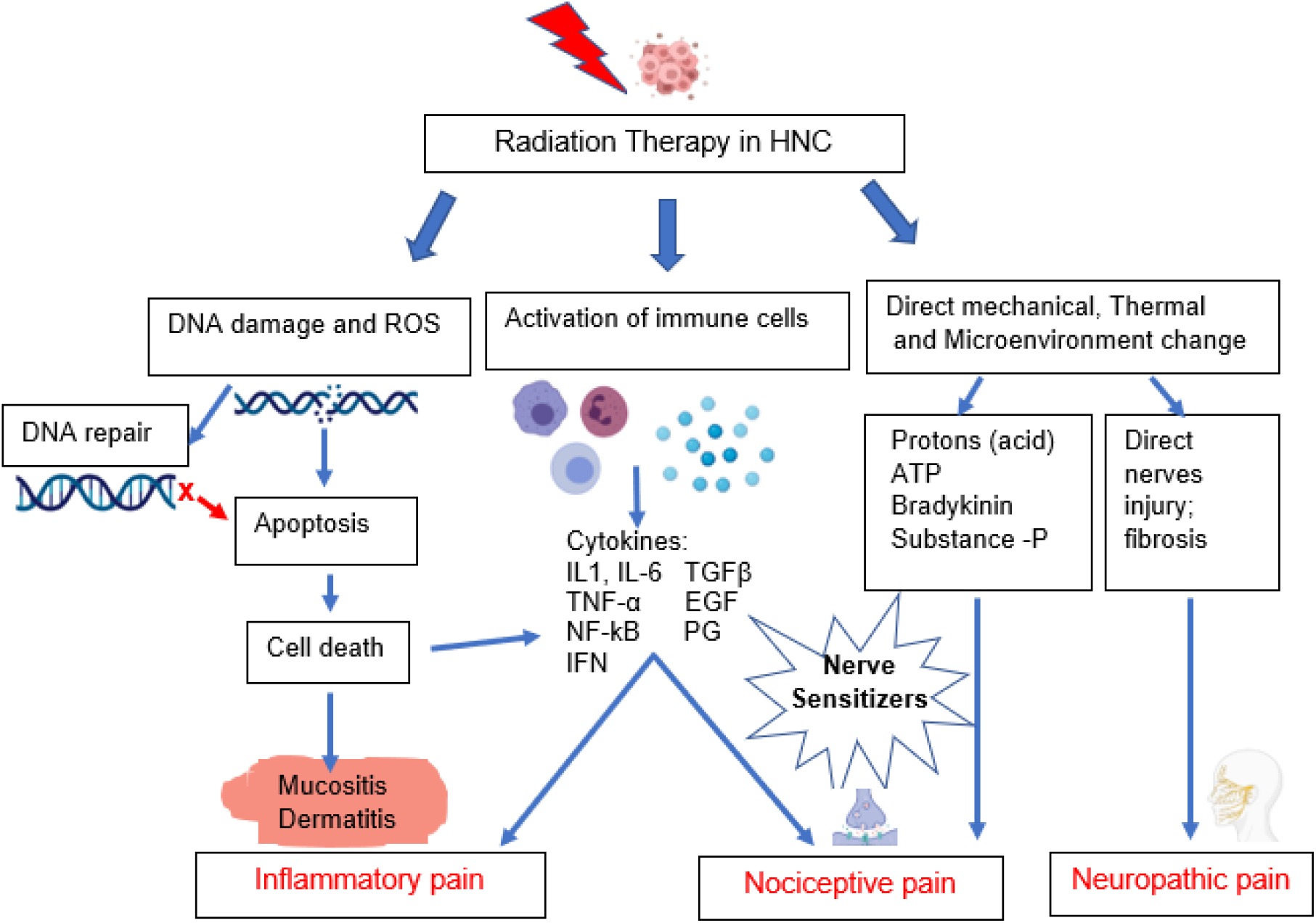
Types and potential mechanisms of radiotherapy induced acute pain. **1-** Radiotherapy in head and neck cancer induces DNA damage of cancer cells and normal neighbor cells and release of reactive oxygen species (ROS). DNA damage induces activation of DNA repair mechanisms which is impaired in cancer cells. Impairment of repair of damaged DNA induces activation of cell death through stimulation of apoptosis processes. Normal tissue death induces mucositis and dermatitis causing **inflammatory pain. 2-** Radiotherapy induces activation of immune responses and inflammatory cells which release inflammatory mediators such as cytokines and chemokines inducing inflammatory pain. Furthermore, these inflammatory mediators act as noxious stimuli, stimulating nociceptors to induce **nociceptive pain. 3-** Radiotherapy induces direct mechanical and thermal damage in addition of change in the microenvironment creating an acidic pH and increase in the adenosine triphosphate (ATP), and release of bradykinin and substance P which act as nociceptors stimuli inducing nociceptive pain. Radiotherapy induces direct nerve injury and inflammation of peripheral nervous system, and fibrosis in the connective tissue can also induce **neuropathic pain**.

**Figure 2:**
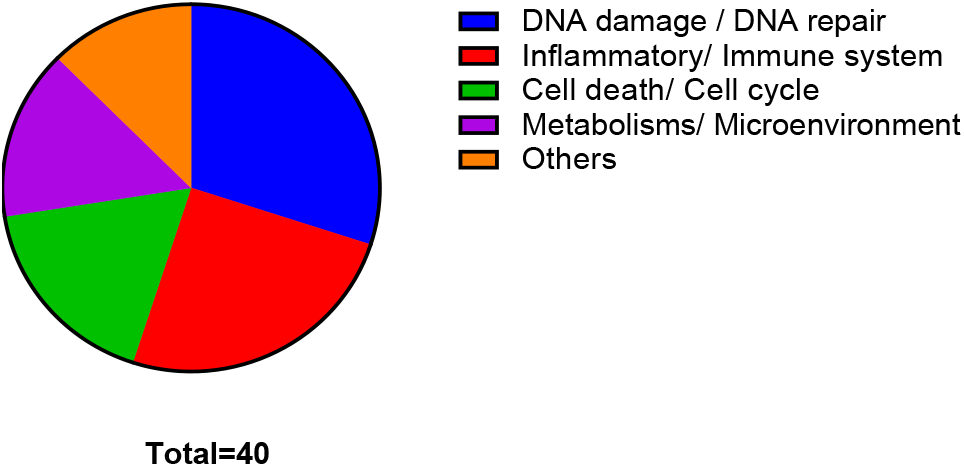
Pie chart of genes associated with radiotherapy induced inflammatory pain; showing the frequency of genes involved different pathways.

### 2 Variants associated with radiotherapy induced Neuropathic pain

Neuropathic pain is one of the common types of pain developed after receiving RT in HNC. Few studies focused on identification of SNPs associated with radiotherapy-induced neuropathic pain. Reyes-Gibby et.al study was the only literature published to identify SNPs associated with neuropathy and neuropathic pain in HNC. 4 variants in 4 genes were identified, associated with radiotherapy induced neuropathic pain. (Table 3)

**Table 3:** Summary of literature search of variants associated with radiotherapy induced acute neuropathic pain

| SNP | Gene | Pathway | Phenotype | Sample/cancer | Number of patients | Author /year |
| --- | --- | --- | --- | --- | --- | --- |
| **rs10950641** | SNX8 | Endocytosis, endosomal sorting and signaling. Glutamatergic-receptor-dependent neural plasticity | Neuropathic pain | DNA/ HNC | 1,043 | Reyes-Gibby et.al. / 2018 <sup>92</sup> |
| **rs6796803** | KNG1 | kinins act as mediators of pain and inflammation | Neuropathic pain | DNA/ HNC | 1,043 | Reyes-Gibby et.al/2018 <sup>92</sup> |
| **rs4775319** | RORA | Expression pattern in lamina II of SC, a region for relaying somatosensory signals of touch, temperature and pain | Neuropathic pain | DNA/ HNC | 1,043 | Reyes-Gibby et.al. / 2018 <sup>92</sup> |
| **rs4804217** | PCP2 | Neuron-specific modulator of intracellular signaling via G proteins | Neuropathic pain | DNA/ HNC | 1,043 | Reyes-Gibby et.al. / 2018 <sup>92</sup> |

### 3 Variants associated with mixed types of radiotherapy induced acute pain

Our literature search revealed some studies identified SNPs associated with post RT-pain without classification of type of pain, we considered this phenotype as mixed types of pain. 13 variants in 10 genes associated with RT-induced Throat/Neck pain and other mixed types of post RT-pain as main phenotype (Table 4).3 genes (21.4%) are involved in cell death and apoptosis pathways, 3 genes (21.4%) are involved in metabolic and microenvironment processes; EGFL6 gene plays a role in angiogenesis ^93^, aquaporin gene AQP7 which plays a rule in water transport through water-selective membrane channel ^94^, and PLAUR gene which plays a role in proteolysis-independent signal transduction ^95^. 1 gene are enriched in neurotransmission/neuropathy pathways or play roles as nociceptors stimuli,1 gene (1%) has a function in DNA damage responses pathway; ATM gene ^96^. (Figure 3, Table 5).

**Table 4:**
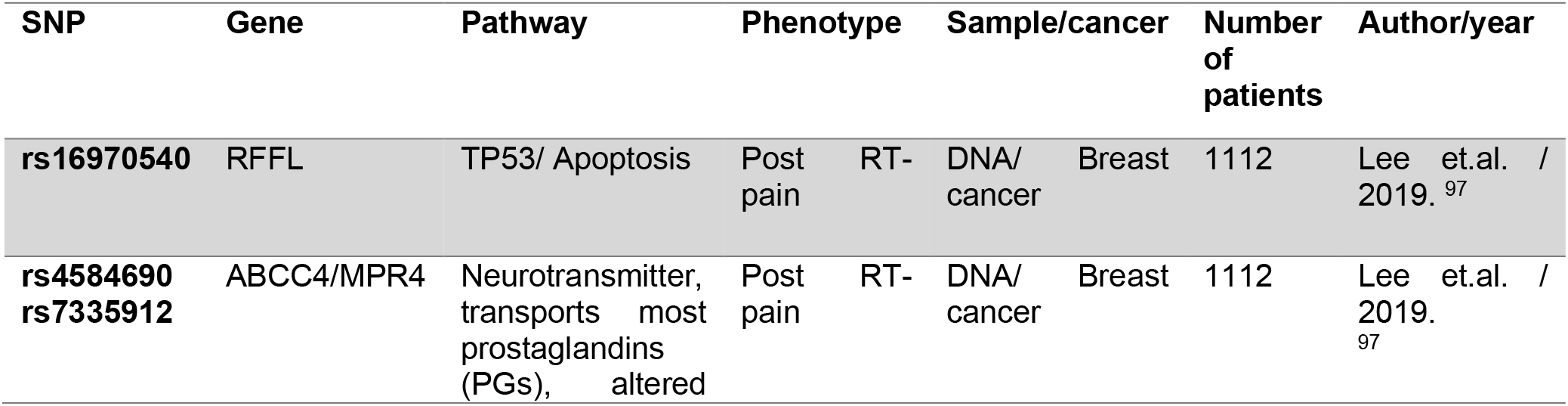
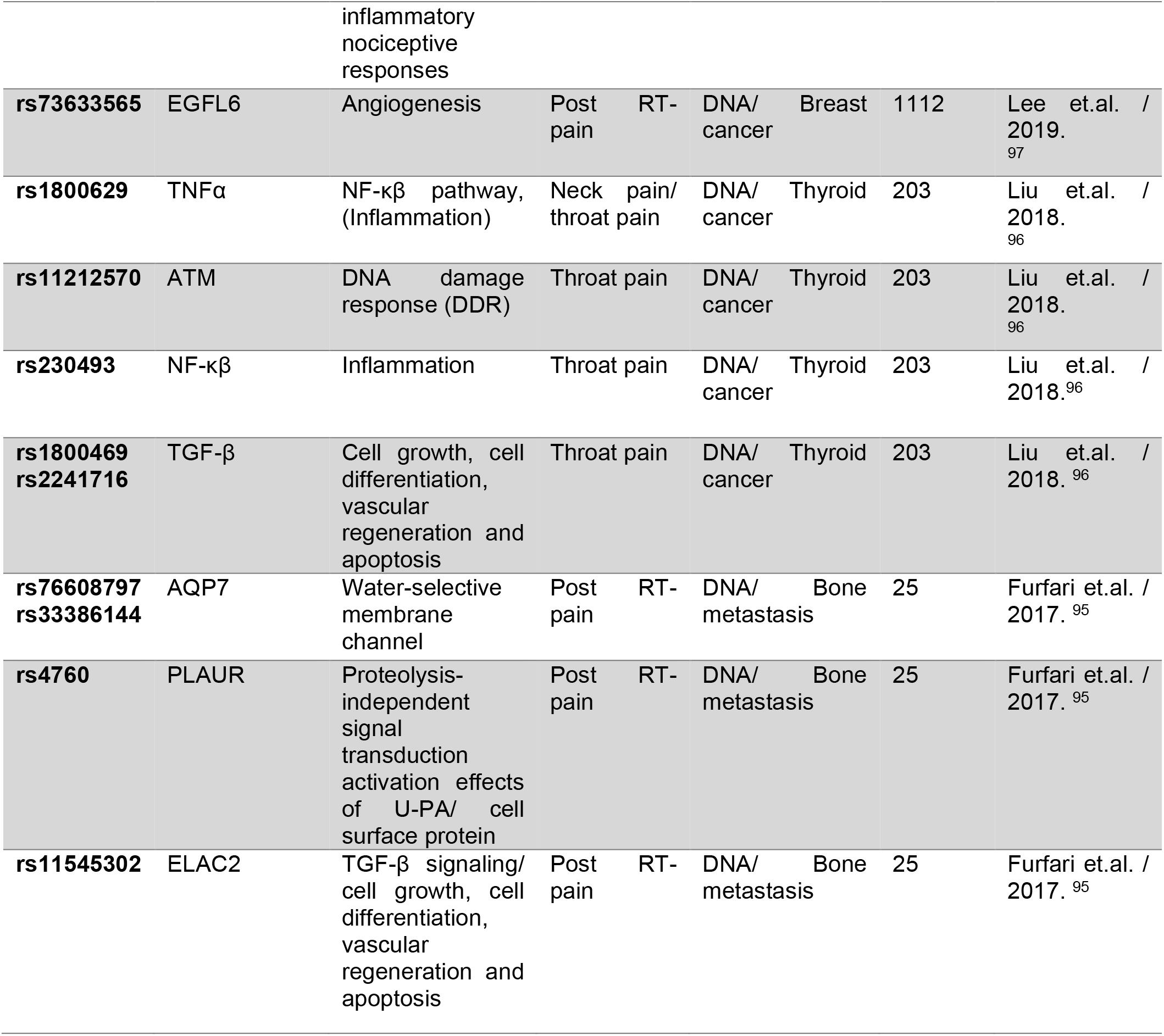
Summery of literature search of variants associated with other types of radiotherapy induced acute pain.

**Table 5:**
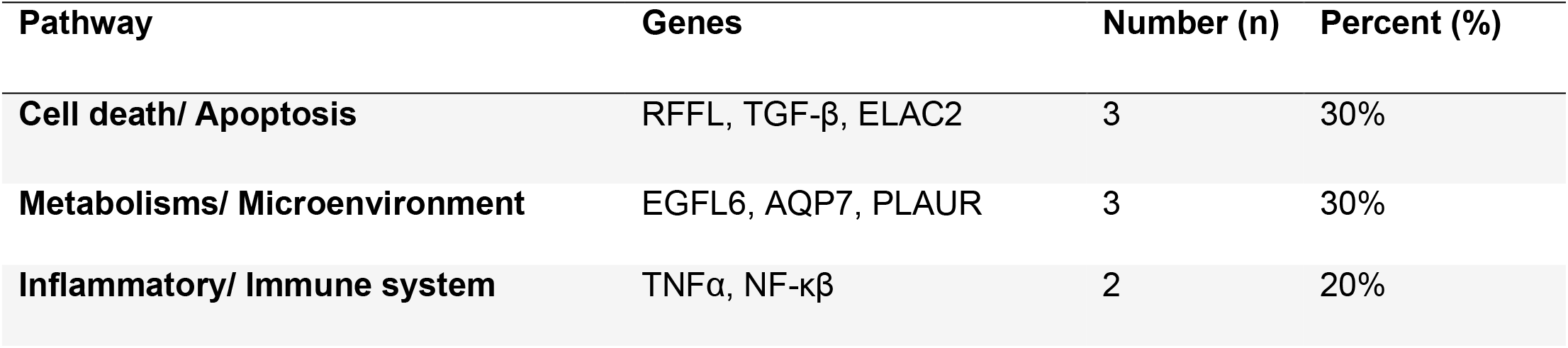
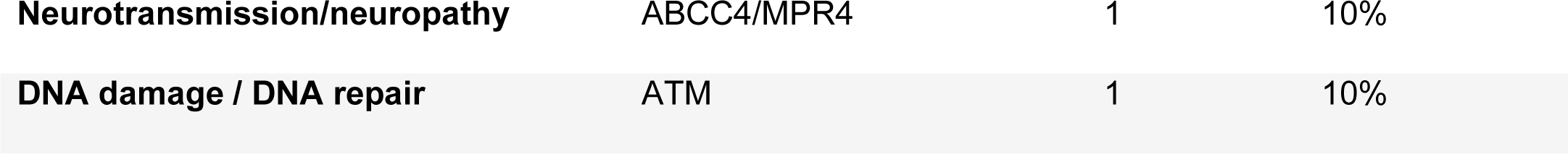
Pathways and biological functions of genes associated with mixed types of radiotherapy induced acute pain.

| Pathway | Genes | Number (n) | Percent (%) |
| --- | --- | --- | --- |
| **Cell death/ Apoptosis** | RFLL, TGF-β, ELAC2 | 3 | 30% |
| **Metabolisms/ Microenvironment** | EGFL6, AQP7, PLAUR | 3 | 30% |
| **Inflammatory/ Immune system** | TNFα, NF-κB | 2 | 20% |
| **Neurotransmission/neuropathy** | ABCC4/MPR4 | 1 | 10% |
| **DNA damage / DNA repair** | ATM | 1 | 10% |

**Figure 3:**
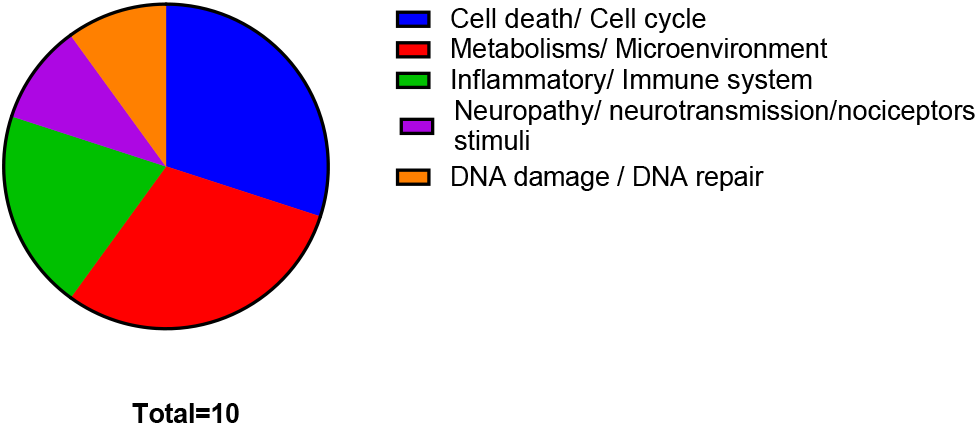
Pie chart of genes associated with radiotherapy-induced mixed types of pain; showing the frequency of genes involved different pathways.

## Discussion

Acute pain is a significant toxicity during and after radiotherapy in HNC, resulting in increase in disability risks, increase in morbidity and decrease in the quality of life of these patients. Acute pain control resulting from RT is highly challenging due to the multifactorial clinical, genetic and molecular mechanisms. Understanding the potential mechanisms and the host genetic variability can help in pain prediction, prevention and in decision making for personalized pain management. To date, few studies explore the mechanisms of acute pain in HNC patients receiving RT. Moreover, very few variants have been identified associated with RT-induced acute pain and relevant acute toxicities in HNC. In this systematic review, we studied the different potential mechanisms for acute pain developed during RT in HNC cancer patients.SNPs identified associated with RT-induced acute pain, mucositis and dermatitis in different types of cancers were collected for comprehensive search.

Our review found that DNA damage, DNA repair, cell death and inflammation are essential in inducing inflammatory pain following RT. Variants detected in genes involved in DNA damage response or DNA repair are associated with acute toxicities (mucositis and dermatitis) related to RT. 18 published studies evaluated the association of variants in DNA damage and DNA repair with RT-induced acute toxicities, 6 out of the 18 studies focused on variants in X-Ray Repair Cross Complementing 1 gene (XRCC1) as a DNA repair gene highly associated with radiotherapy toxicity ^61, 63, 74, 75, 98^. Other variants in DNA damage and repair genes; XRCC2, XPD, ERCC1, ZNF24, NBN, Ku70, ATM, CHEK1, RAD51C, ERCC2, PMS1, MLH1 were associated with RT-acute toxicities, these results matched our study of the potential mechanisms of inflammatory pain induced by RT ^22^. Additionally, 11 studies identified variants in genes involved in inflammatory pathways and immune systems including; TNF-α ^60, 96, 99^, STING1 ^100^, HLA-DQB1^58^, TNFRSF1A ^64^, GHRL ^65^, PLA2G2F/ IIF ^101^, IFNG ^80^, IL12RB2 ^91^ and NF-κβ ^96^, these variants were associated with RT-induced inflammatory pain and mixed throat pain. Studies showed that activation of inflammatory and immune cells after RT induces release of inflammatory cytokines and chemokines promoting more cell damage inducing inflammatory pain in addition to activation of nociceptors causing nociceptive pain ^32, 33, 102–104^. Reyes-Gibby et.al, studied the rule of cytokines in pain activation and sensitization ^53, 105^. Variants in genes regulating release of pro-inflammatory mediators as cytokines and chemokines are shown here in our review.

Few studies focused on neuropathic pain induced by RT, Reyes-Gibby et.al identified 4 SNPs in 4 genes (SNX8, PCP2, RORA and KNG1) correlated with neuropathy in HNC ^92^. Although substance-P and other neurotransmitters as excitatory amino acids exaggerates pain response ^53, 106^, no study have identified any variant in Substance-P or other neurotransmitters amino acids correlated with RT-induced pain, more studies are needed to identify the SNPs related to RT-induced neuropathy and neuropathic pain, specially variants in genes involved in neuronal plasticity and neuroinflammatory pathways. Other cell signaling pathways including cell cycle and NF-kB pathway (CCND1) ^67, 68^, autophagy (ATG16L2, ATG10) ^62^, wnt/β-catenin (APC, GSK3β) ^71^ and angiogenesis (EDN1)^69^ regulating genes were associated with RT related acute mucositis and dermatitis.

Studies included in this review, used either candidate gene approach or genome wide association analysis approach (GWAS), and interestingly through GWAS studies collected, we declared variants in genes involved in different/new pathways other than the common potential mechanisms of RT-induced pain, associated with RT-induced acute pain and toxicities. Sckack et.al conducted GWAS in HNC patients and identified candidate (rs1131769) in DNAJC18 gene regulating a chaperon molecules protecting cellular proteins, significantly associated with oral mucositis (OM) after RT ^57^. Yang et.al did GWAS of NPC patients and identified variant (rs117157809) in Tankyrase (TNKS) gene regulating telomere capping and maintenance of telomerase activity, is significantly associated with RT-induced OM ^59^. Li et.al, identified GWAS risk loci (rs1265081) in CCHCR1 gene regulating cellular process as keratinocyte proliferation, differentiation and EGFR pathways, associated with RT-induced OM in HNC patients ^58^, while Rattay et.al, identified variant (rs12554549) in PTPN gene regulating cell growth, differentiation and oncogenic transformation, associated with RT-induced acute skin ulceration in breast cancer patients ^78^. Genetic variants involved in oxidative stress response and ROS metabolism pathways (e.g. GSTM1 and NOS3) were associated with RT acute skin toxicity in breast cancer patients ^81, 107^

Although we did a comprehensive review to identify different potential mechanisms, including molecular pathways and genetic variants associated with RT-induced acute pain and relevant acute toxicities, there are limitations in our review. such as we focused on the most common acute toxicities being the common risk of acute pain as oral mucositis and dermatitis, while there are other acute toxicities could be relevant such as dysphagia. We did a comprehensive search covering all types of cancers, since studies focused on HNC were few and we would like to highlight other variants detected in other cancer types to be validated and focused on in future studies. Furthermore, many studies didn’t specify the type of pain detected either inflammatory, nociceptive or neuropathic, while they only mentioned post-RT pain or throat pain, we involved these variants under the group of other types/mixed pain. These studies collected, are just association studies, however, mechanistic studies must be done to test the mechanistic effect of these variants on the phenotype development and alteration of the different pathways associated. These identified candidates need further validation through more studies replication. More comprehensive analysis of the other candidates relevant to different types pf pain not associated with cancer or RT, is considered in our future plans.

Our future directions include validating these genetic variants and developing a predictive algorithm for pain prediction and management optimization using genetic candidates identified in out HNC cohort.

## Conclusion

Multiple studies were conducted to identify different variants associated with cancer pain and cancer treatment induced pain. However, few of them focused on radiotherapy induced acute pain which is considered almost the most common acute toxicity following RT. We did a comprehensive literature review to identify the variants and genes that have been previously reported to have association with radiotherapy induced acute pain phenotypes including; mucositis, dermatitis (Inflammatory pain), neuropathic, nociceptive pain and mixed oral/mouth pain. Our review revealed that pain is a complex symptom following RT in OC/OPC patients due to its multifactorial origin and different phenotypes of its representation. DNA damage/ repair, inflammatory pathways, apoptosis, cell death, and neuropathy pathways are the most common pathways behind the development of radiotherapy-related acute toxicities and acute pain. Furthermore, there is a need for a comprehensive understanding of the genetic profile in addition to the clinical and treatment background of those patients who have high risk to develop acute pain following RT and to identify a more standardized algorithm for personalized management of acute RT-induced pain to maintain pain relief during RT and improve patients’ outcome and quality of life.

## Data Availability

All data produced in the present study are available upon reasonable request to the authors

## Abbreviations

HNSCC: Head and neck squamous cell carcinoma
OC/OPC: Oral cavity and oropharyngeal cancer
HPV: Human papilloma virus
RT: Radiotherapy
QOL: Quality of life
SSB: Single strand breaks
DSB: Double strand breaks
ROS: Reactive oxygen species
HNC: Head and neck cancer
NPC: Nasopharyngeal cancer
IL: Interleukins
PG: Prostaglandins
TNF: Tumor necrosis factors
Gy: Gray
VEGF: vascular endothelial growth factor signaling
GABA: γ-aminobutyric acid
NSCLC: Non-Small Cell Lung Cancer
NGF: Nerve growth factor
ATP: Adenosine triphosphate
SNPs: Single nucleotide polymorphisms
DDR: DNA damage response

