## Supplemental tables for "Systematic Review of Genetic Polymorphisms Associated with Acute Pain and Relevant Acute Toxicities Induced by Radiotherapy for Head and Neck Cancer"

**Supplementary Table S1. Ovid MEDLINE search strategy**

| # | Search |
|---|---|
| 1 | exp Radiotherapy/ |
| 2 | (radiotherap* or irradiat* or radiat* or chemoradi* or radiochemo* or chemo-radi* or radio-chemo* or "intensity modulated" or IMRT or EBRT or WBRT or hWBRT).ti,ab,kf. |
| 3 | (proton* adj3 (beam or therap* or radiation*)).ti,ab,kf. |
| 4 | exp Radiotherapy Planning, Computer-Assisted/ |
| 5 | exp Radiation Oncology/ |
| 6 | or/1-5 |
| 7 | limit 6 to english language |
| 8 | (animals not (humans and animals)).sh. |
| 9 | 7 not 8 |
| 10 | (mice or mouse or murine or rat or rats or rodent or cells or "in vitro" or "cell line").ti. |
| 11 | 9 not 10 |
| 12 | exp Pain/ |
| 13 | pain.ti,ab,kf. |
| 14 | Mucositis/ or (mucositides or Mucositis).ti,kf. |
| 15 | exp Dermatitis/ or (dermatitis or dermatitides).ti,kf. |
| 16 | (neuropath* or polyneuropath* or paresthesia or neuralgia* or allodynia* or hyperalgesia*).ti,kf. |
| 17 | exp Analgesics, Opioid/ or (analgesic* or opioid*).ti,kf. |
| 18 | "acute toxicit*".ti,kf. |
| 19 | or/12-18 |
| 20 | 11 and 19 |
| 21 | exp Polymorphism, Single Nucleotide/ |
| 22 | exp Genetic Variation/ |
| 23 | exp Genetic Predisposition to Disease/ |
| 24 | exp DNA Mutational Analysis/ |
| 25 | exp Genome-Wide Association Study/ |
| 26 | exp Genetic Markers/ |
| 27 | exp Pharmacogenetics/ |
| 28 | exp Biomarkers, Tumor/ |
| 29 | exp High-Throughput Nucleotide Sequencing/ |
| 30 | exp Mutation/ |
| 31 | exp DNA Repair/ |
| 32 | exp gene expression regulation/ |
| 33 | exp gene expression/ |
| 34 | ((genetic or molecular or "single nucleotide") adj3 (biomarker* or marker* or variation* or variant* or test*)).ti,kf. |
| 35 | (pharmacogenomic* or pharmacogenetic* or polymorphism* or SNP* or mutation* or "gene expression").ti,kf. |
| 36 | exp Molecular Diagnostic Techniques/ |

| | |
|---|---|
| 37 | exp Genetic Techniques/ |
| 38 | exp DNA, Neoplasm/ |
| 39 | Genetics/ |
| 40 | Genetics.fs. |
| 41 | or/21-40 |
| 42 | 20 and 22 |

**Supplementary Table S2. Ovid Embase search strategy**

| # | Search |
|---|---|
| 1 | exp radiotherapy/ |
| 2 | (radiotherap* or irradiat* or radiat* or chemoradi* or radiochemo* or chemo-radi* or radio-chemo* or "intensity modulated" or IMRT or EBRT or WBRT or hWBRT).ti. |
| 3 | exp radiotherapy planning system/ |
| 4 | 1 or 2 or 3 |
| 5 | limit 4 to english language |
| 6 | Human/ |
| 7 | Nonhuman/ or ANIMAL/ or Animal Experiment/ |
| 8 | 7 not 6 |
| 9 | 5 not 8 |
| 10 | (mice or mouse or murine or rat or rats or rodent or cells or "in vitro" or "cell line").ti. |
| 11 | 9 not 10 |
| 12 | pain.ti,ab,kf. |
| 13 | exp mucosa inflammation/ |
| 14 | exp mucosa inflammation/ or (mucositides or Mucositis).ti,kf. |
| 15 | exp dermatitis/ |
| 16 | exp dermatitis/ or (dermatitis or dermatitides).ti,kf. |
| 17 | (neuropath* or polyneuropath* or paresthesia or neuralgia* or allodynia* or hyperalgesia*).ti,kf. |
| 18 | exp narcotic analgesic agent/ or (analgesic* or opioid*).ti,kf. |
| 19 | "acute toxicit*".ti,kf. |
| 20 | or/12-19 |
| 21 | 11 and 20 |
| 22 | exp single nucleotide polymorphism/ |
| 23 | exp genetic variation/ |
| 24 | genetic variability/ |
| 25 | exp genetic predisposition/ |
| 26 | exp pharmacogenetics/ |
| 27 | (genetic adj3 (variation* or variant*)).ti,ab,kf. |

| | |
|---|---|
| 28 | (pharmacogenomic* or pharmacogenetic* or polymorphism* or SNP* or "single nucleotide ").ti,ab,kf. |
| 29 | or/22-28 |
| 30 | 21 and 29 |
| 31 | 2 and 30 |
| 32 | conference abstract.pt. |
| 33 | 31 not 32 |

**Supplementary Table S3. Clarivate Analytics Web of Science search strategy**

| | |
|---|---|
| 1 | TS=(radiotherap* or irradiat* or radiat* or chemoradi* or radiochemo* or "chemo-radi*" or "radio-chemo*" or "intensity modulated" or IMRT or EBRT or WBRT) |
| 2 | (mice or mouse or murine or rat or rats or rodent or cells or "in vitro" or "cell line") (Title) |
| 3 | #1 NOT #2 |
| 4 | TS=(pain or mucositides or Mucositis or dermatitis or dermatitides or neuropath* or polyneuropath* or paresthesia or neuralgia* or allodynia* or hyperalgesia* or analgesic* or opioid* or "acute toxicit*") |
| 5 | #3 AND #4 |
| 6 | TS=("single nucleotide" or polymorphism* or SNP* or "genetic variation*" or "genetic variant*" or “genetic predisposition*” or pharmacogenetics or pharmacogenomic*) |
| 7 | #5 AND #6 and Meeting Abstracts (Exclude – Document Types) |
| 8 | #5 AND #6 and Meeting Abstracts (Exclude – Document Types) and English (Languages) |
